# Sensory evaluations of a novel iron and zinc-enriched powder for the potential treatment and prevention of iron deficiency in women of reproductive age

**DOI:** 10.1101/2022.12.10.22283294

**Authors:** Clara H Miller, Hauna Sheyholislami, Jessie L Burns, Kristin L Connor

## Abstract

Iron deficiency (ID) and iron deficiency with anaemia (IDA) are serious global health problems that disproportionately affect women aged 15-49 years. Although food fortification is one of the most effective and sustainable ways to combat nutritional deficiencies, iron remains one of the most difficult micronutrients to fortify, given its tendency to react strongly with food constituents. Therefore, it is important to assess the sensory properties of foods fortified with iron to determine the acceptability and palatability in target populations. We aimed to determine the palatability and acceptability of a novel iron and zinc enriched powder fortified in tap water by conducting sensory evaluations in 35 women of reproductive age using a 9-point hedonic scale, where participants rated the sensory properties of six samples containing different amounts of the active or placebo. We found significant differences between samples reconstituted at 1g/L, 2g/L, and 3g/L for sensory properties, including overall taste. Participants were found to be more willing to drink the mineral-enriched powder when prepared at the lowest concentration (1g/L) compared to higher concentrations. Our results provide important insight on sensory qualities of a novel formulation of an iron and zinc -enriched powder for at-home fortification, and indicate consumer acceptability in reproductive aged women, a key group at risk for ID/IDA. If found to improve iron status, novel treatments like this product will contribute to global efforts to develop safe, acceptable and sustainable interventions for ID and IDA.

## Introduction

Iron and zinc are two of the most common mineral deficiencies worldwide ^1,2^. These deficiencies are often caused by inadequate dietary intakes of these minerals and can result in serious adverse health problems if left untreated ^3,4^. Both iron and zinc are important for proper immune function and disease resistance ^5,6^. Iron and zinc deficiencies often occur simultaneously as these nutrients are frequently found in many of the same foods, and their absorption in the body is affected by many of the same dietary ligands ^7^. Notably, zinc also serves as an important catalyst in iron metabolism, meaning that low dietary intake of zinc can contribute to the development of iron deficiency (ID)^8,9^.

ID affects approximately 20-25% of the population and 52% of pregnant people ^10-12^. Women of reproductive age (15-49 years) have an increased risk of developing ID due to their increased physiologic demand for iron as a result of menstruation and pregnancy^4,13^. ID results in fatigue, weakness, cognitive dysfunction, and decreased immunity, which ultimately affects quality of life ^1,4,13,14^. If left untreated,

ID can develop into iron deficiency anaemia (IDA), which is the most common form of anaemia globally ^11^. IDA exacerbates the symptoms of ID while introducing additional health risks, such as cardiac complications ^15^ due to reduced erythropoiesis, which, in pregnant people, threatens maternal health and birth outcomes ^14,16,17^. Currently, IDA is a serious public health problem affecting one in three women aged 15-49 worldwide, and 10% of women aged 15-49 in Canada ^11,18,19 20^. Therefore, there is an urgent need to ensure adequate iron status in women of reproductive age to prevent adverse health problems and ultimately promote healthy pregnancies.

According to the Canadian Clinical Guidelines for Family Medicine ^21^, the first-line treatment for ID and IDA in most patients is oral iron-replacement therapy, in addition to dietary modifications ^21,22^. Current interventions include supplementation with iron salts (such as ferrous gluconate, ferrous fumarate, and ferrous sulphate) and other at-home fortification systems ^23-25^. However, many of these interventions have only been moderately successful due to low long-term (more than 18-months) compliance (less than 50%), which decreases the overall effectiveness of the intervention^26-28^.

Low compliance rates with current treatments are mainly attributed to unpleasant side effects, including nausea and constipation (associated with iron supplementation)^29-31^. Iron fortification is more gentle on the digestive system than supplementation with iron salts ^29,32^, resulting in fewer side effects (such as nausea, flatulence, abdominal pain, constipation, and black or tarry stools) ^29,30,32,33^. However, fortification with iron has historically been known to affect various sensory modalities (taste, colour, odour) when reconstituted in food and drink, causing poor palatability, and ultimately resulting in low consumer compliance rates ^27,34^.

To more effectively prevent and treat ID and IDA in women of reproductive age, new interventions with improved consumer adherence are required. Such interventions would contribute to sustainable solutions to address these conditions and improve the health of populations globally. To meet this need, we investigated the palatability and acceptability of an iron and zinc-enriched powder composed of a novel formulation of electrolytic ferrous iron with a small particle size (<20 μm) and zinc sulphate monohydrate that can be reconstituted in tap water for at-home fortification. The main purpose of this intervention is to address iron deficiency, however, the formulation of the enriched powder also contains zinc, given that low baseline zinc levels can negatively affect iron absorption^9^. Further, iron and zinc deficiencies often occur simultaneously due to these minerals being frequently found in many of the same foods and populations who are affected by malnutrition may not be consuming adequate amounts of foods containing these minerals^35,36^. To establish regulatory guidelines for this product, it is important to demonstrate that fortification with the mineral-enriched powder is palatable to end-users when reconstituted in tap water. This powder has been designed to affect the sensory properties of the water (odour, colour, and taste) only minimally in order to circumvent the problem of poor palatability. An at-home fortification system for iron that is palatable would suggest good consumer acceptability and may ultimately contribute to higher compliance rates ^23,34,37-40^. Therefore, the objectives of our study were to determine whether the mineral-enriched powder affects sensory perceptions (odour, colour, and taste) when reconstituted in tap water and if the reconstituted solution is palatable and acceptable to women of reproductive age.

## Methods

### Study Design and Population

This study aimed to determine the palatability and acceptability of an iron and zinc micronutrient-enriched powder compared to a placebo powder. The study was conducted as a single-blinded study, where only the researcher knew which samples contained the intervention and which samples contained the placebo, and their concentrations, between November 2021 and March 2022 in Ottawa, Ontario, Canada. Participants were biologically female women of reproductive age (18-35 years), a target consumer population for this intervention. All participants were recruited locally from a university campus. This study was approved by Office of Research Ethics at Carleton University (CUREB-B Protocol #116204).

Interested participants met virtually with a member of the research team for a 5-minute phone or Zoom call to discuss the objectives of the study and review the screening criteria. Eligible participants were those who were: women of the biological female sex (assigned at birth), between 18-35 years old, and not currently taking any mineral supplements. Only participants who were not taking any supplements containing minerals were included in this study because some nutritional supplements contain iron, chromium, calcium, and zinc which can cause temporary changes in taste perception ^41,42^.

Fifty-two participants were screened for this study. Of these, 35 were deemed eligible and agreed to participate in the study (Figure 1).

**Figure 1.**
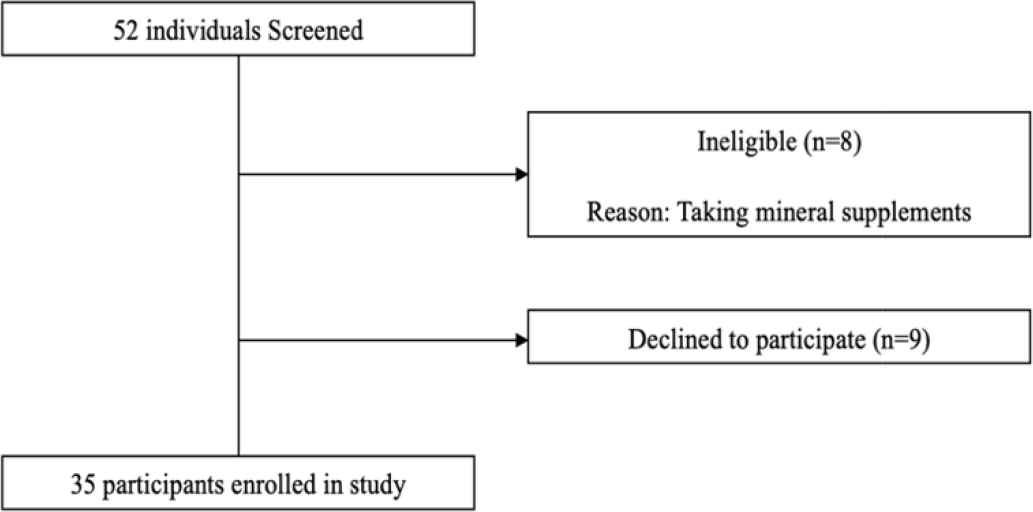
Participant recruitment and enrolment flow-chart.

### Study Products

This study sought to evaluate the palatability and acceptability of three different concentrations of a mineral-enriched powder dissolved in water, compared to a placebo. Participants evaluated a total of six samples, three samples containing the intervention and three containing a placebo. The three samples containing the intervention were grouped as the “active samples”. These samples were labelled S1, S2, S3 and contained 1g/L, 2g/L, and 3g/L of the mineral-enriched powder, respectively. The three placebo samples were labelled S4, S5, S6 and contained 1g/L, 2g/L, and 3g/L of the placebo powder, respectively.

The iron and zinc enriched powder has been developed using a fine-grade electrolytic iron powder that has been approved for use and regularly incorporated into breakfast cereals worldwide. The zinc is derived from zinc sulphate monohydrate and is approved for supplementation. These minerals are included in a proprietary mixture of food stabilisers (primarily vegetable-derived gums) and a milled vinegar, adapted by the manufacturer (Gum Products International [GPI] Inc.). The placebo powder was composed of the same proprietary mixture as the active powder, only without the added iron and zinc. All constituents of the mixture are designated by Health Canada as safe for human consumption and meet the international Food Codex standards for food additives ^43^. The levels of iron and zinc in the formulation are at levels similar to those used for routine fortification of staple products such as flour, corn flakes and other cereals, and well below the levels where toxicity would be a concern.

### Evaluation of Palatability and Acceptability of Study Products

Eligible individuals were invited to individually participate in a 20-30-minute sensory evaluation session conducted in-person. Only one participant and one member of the research team were present during each session. During the study, participants were presented with a total of six samples to evaluate: three samples containing the intervention and three samples containing a placebo. Each sample was prepared by dissolving different amount of either the mineral-enriched powder or the placebo powder in 1L of water.

Each participant was presented with the six samples to evaluate in a random order, given a score card, and provided with instructions on how to perform the sensory tests (for odour, colour, and taste). The score cards were used to capture information about the palatability of the active and placebo preparations, using a 9-point hedonic scale ^44^.

The 9-point hedonic scale measures how much an individual likes or dislikes a food product ^44^. The scale ranges from 1-9, with a score of 9 meaning “like extremely” and a score of 1 meaning “dislike extremely”. A score of 5 indicates a neutral response, meaning “neither like nor dislike”. These hedonic scores are interpreted to understand the palatability of the samples: A score of 5 indicates that the sensory property is palatable, a hedonic score of 7 or greater indicates that the sensory property is well liked (good acceptability by the consumer ^44-46^) and a hedonic score of 4 or less indicates that the sensory property is disliked (and not acceptable to the consumer ^44-46^).

Participants were asked to use the 9-point hedonic scale to evaluate the sensory properties of colour, odour, overall taste, and the 5 organoleptic properties of taste (sour, sweet, bitter, salty, and umami) for each sample by recording their answers on the provided score card. Participants were also asked “*would you be willing to drink this sample daily?*” to determine the acceptability of the samples.

### Statistical Analysis

Within groups (active and placebo), hedonic scores for each sensory property (odour, colour, salty, sweet, bitter, sour, and umami) and overall taste were compared between concentrations (1 g/L, 2 g/L, and 3 g/L) using the Friedman’s two-way analysis of variance by ranks test. When there were significant differences found between the three groups, Wilcoxon signed-rank tests were performed to determine individual differences between groups, and p-values were adjusted for multiple comparisons with Bonferroni correction. Hedonic scores were also compared between groups (active vs placebo) at each concentration using the Wilcoxon signed-rank test and visualized in polar plots. Responses to the question “*would you be willing to drink this sample daily?*” (yes/no) were reported as the proportion of participants who answered “yes” or “no” to this question (95% confidence interval).. Data were analysed using RStudio Statistical Software (v2022.02.1+461). Hedonic scores for the five organoleptic properties of taste are reported as medians and interquartile ranges (IQR) and visualized using polar plots (RStudio version 2022.02.1+461). All values were considered to be statistically significant at p<0.05.

## Results

### Mineral-enriched samples score higher than placebo samples on the hedonic scale

Overall, with increasing concentration of the active and placebo powders, hedonic scores decreased (Figure 2). At the lowest concentration (1 g/L), there were significant differences between the active intervention and placebo samples for the sensory properties of “colour” (p<0.001), “salty” (p=0.01), “sweet” (p=0.03) and “sour” (p=0.03) (Figure 2). Participants also rated the “overall taste” of the active powder at 1 g/L (S1) higher than the placebo at 1 g/L (S4) (p=0.01 Figure 2). At a concentration of 2 g/L, there were significant differences between the active intervention (S2) and the placebo samples (S5) for the sensory properties of “colour” (p<0.001), “salty” (p=0.002), “sweet” (p=0.02), “bitter” (p=0.004), “umami” (p=0.04) and “sour” (p=0.002) and in the “overall taste” (p<0.001, Figure 2). At a concentration of 3 g/L, there were significant differences between the active intervention (S3) and placebo samples (S6) for the sensory properties of “colour” (p<0.001), “odour” (p=0.006), “salty” (p<0.001), “bitter” (p=0.03), “umami” (p=0.02), and “sour” (p=0.01), and in the “overall taste” (p<0.001, Figure 2).

**Figure 2.**
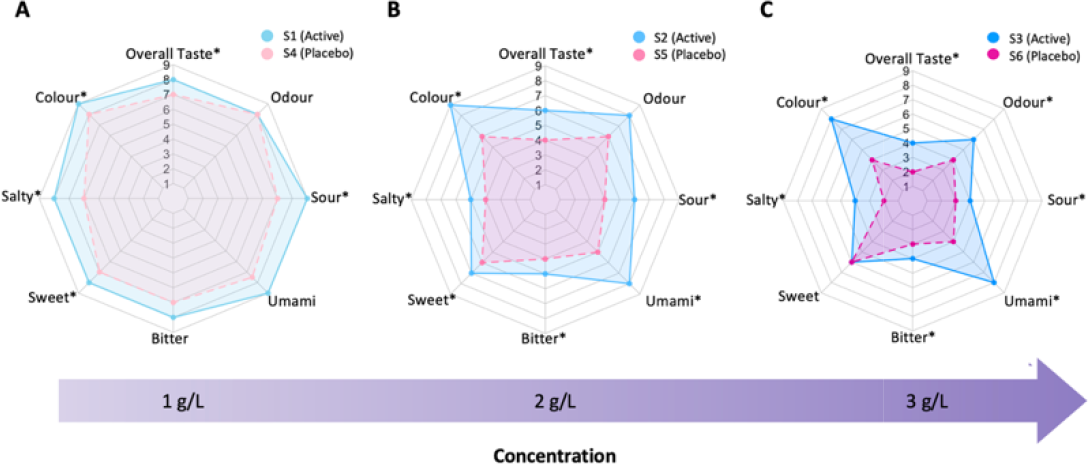
Participant evaluations for seven sensory properties (colour, odour, salty, sweet, bitter, sour, umami) and overall taste for active and placebo solutions at three concentrations. Sensory evaluations were reported using a numeric 9-point hedonic scale. A. Hedonic ratings of 1g/L active and placebo solutions. B. Hedonic ratings of 2g/L active and placebo solutions. C. Hedonic ratings of 3g/L active and placebo solutions. A hedonic score of 7 or greater indicates the sample will likely have good consumer acceptability. A score of 5 or greater indicates the overall taste of the sample (or sensory property) is palatable. S1, S2, S3 (blue) = active solutions containing the mineral-enriched powder at concentrations of 1g/L, 2g/L, and 3g/L, respectively. S4, S5, S6 (pink) = placebo solutions containing the control powder at concentrations of 1g/L, 2g/L, and 3g/L, respectively. * indicates a significant difference between a sensory property in the active vs placebo solution (Wilcoxon signed-rank test, p<0.05).

### Favourable sensory perceptions decrease with increasing concentration of mineral-enriched samples and placebo samples

To determine which concentrations of the active and placebo preparations were more palatable and liked, sensory properties were evaluated across the concentrations within each group (active and placebo). For the active preparations (S1, S2, S3), there were significant differences across concentrations in participants’ reporting of all sensory properties other than “umami” (p<0.05, Supplementary Table 1). The 1 g/L (S1) active sample received the highest hedonic scores for each sensory property compared to the active samples at 2 g/L or 3 g/L (Supplementary Table 1).

Notably, the median hedonic score for the “overall taste” of the active sample at 1 g/L (S1) was also significantly higher when compared to the median hedonic scores for “overall taste” in both the active sample at 2 g/L (S2) (p<0.05, [1.00, 2.99]) and the active sample at 3 g/L (S3) (p<0.001, [3.49, 4.50], Figure 3). Further, to better understand the distribution of scores from participants, the proportion of participants who disliked (hedonic scores 1-4), were neutral towards (hedonic score of 5), or liked (hedonic scores of 6-9) the “overall taste” of each active sample were determined (Supplementary Table 2). In total, 82% of participants indicated that they liked the overall taste of sample S1, 57% indicated that they liked sample S2, and only 6% indicated that they liked sample S3 (Supplementary Table 2).

**Figure 3.**
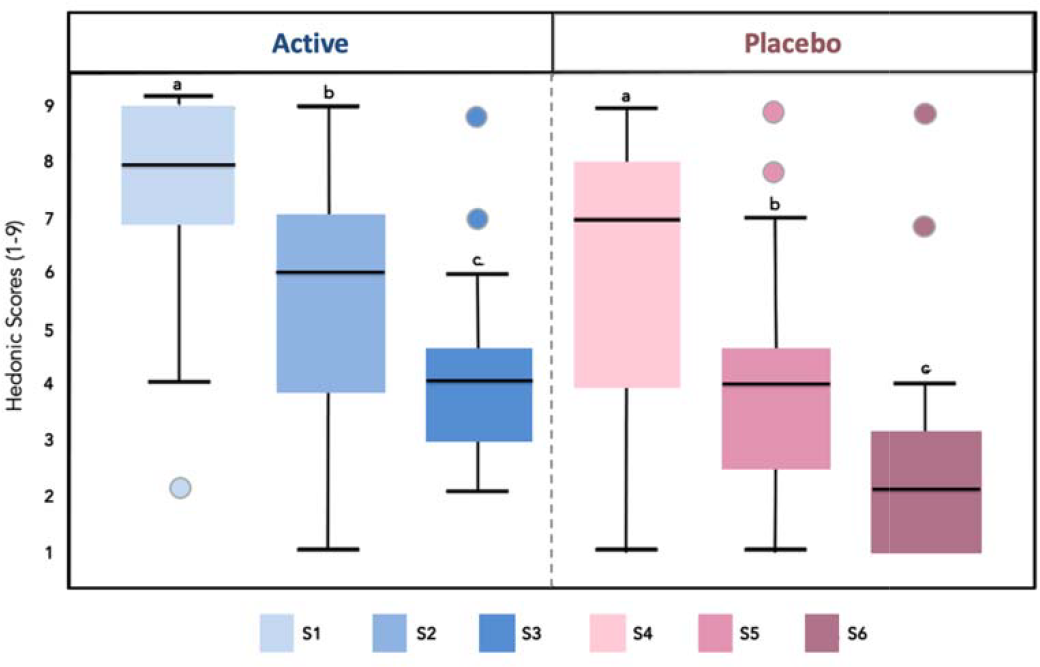
Participant evaluation of the overall taste for active and placebo solutions. S1, S2, S3 (blue) = active solutions containing the mineral-enriched powder at concentrations of 1g/L, 2g/L, and 3g/L, respectively. S4, S5, S6 (pink) = placebo solutions containing the control powder at concentrations of 1g/L, 2g/L, and 3g/L, respectively. Data are median and IQR. Within panels, groups with different letters are significantly different (Wilcoxon signed-rank test with Bonferroni correction for multiple comparisons, p<0.05).

For the placebo preparations (S4, S5, S6), there were significant differences across concentrations in participants’ reporting of six out of the seven sensory properties (“odour”, “colour”, “salty”, “sweet”, “bitter” and “sour”, p<0.001, Supplementary Table 1). The placebo sample at 1 g/L (S4) received the highest hedonic scores for each sensory property compared to samples at 2 g/L or 3 g/L (Supplementary Table 1). The median hedonic score for the “overall taste” of the placebo sample at 1 g/L (S4) was significantly higher when compared to the median hedonic score for both the placebo sample at 2 g/L (S5) (p<0.001, [1.50, 3.50]) and at 3 g/L (S6) (p<0.001, [3.00, 4.99]) (Figure 3). The proportion of participants who disliked, were neutral towards, or liked the “overall” taste of each placebo sample was also determined (Table 2). In total, 60% of participants indicated that they liked the overall taste of S1, 23% indicated that they liked S2, and only 6% indicated that they liked S3 (Supplementary Table 2).

### Acceptability was highest at lowest sample concentrations

Acceptability of the samples was assessed by comparing participant responses within active and placebo groups to the question “*would you be willing to drink this sample daily?”*. For both the active and placebo samples, the proportion of participants willing to drink the sample daily decreased as the concentration increased. Among the active samples, the proportion of participants who were willing to drink the active sample with the lowest concentration at 1g/L (S1) daily was 97% (85 to 99%), whereas 74% were willing to drink the sample daily when prepared at 2 g/L (S2) (58 to 86%), and only 46% were willing to drink the sample daily when prepared at 3 g/L (S3) (30 to 62%) (Figure 4).

**Figure 4.**
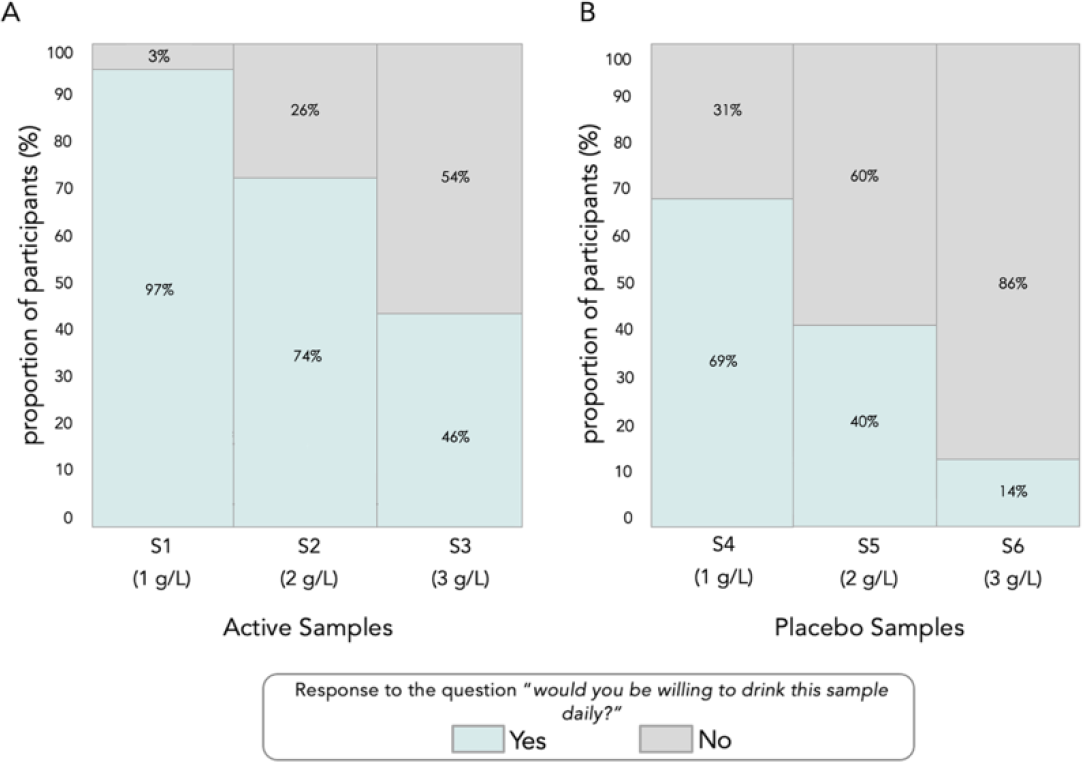
Proportion of participant responses to the question of “*would you be willing to drink this sample daily*?” for each sample (S1-S6). Proportion of participant responses to “yes” (green bars) and “no” (grey bars) at each concentration for active and placebo samples. A. Active samples (S1, S2, S3), containing the mineral-enriched powder. B. Placebo samples (S4, S5, S6), containing the placebo powder.

Among the placebo samples, the proportion of participants willing to drink the sample with the lowest concentration at 1 g/L (S4) daily was 69% (52 to 81%), whereas 40% were willing to drink the placebo sample daily when prepared at 2 g/L (S5) (26 to 56%), and only 14% were willing to drink the sample daily when prepared at 3 g/L (S6) (6 to 29%) (Figure 4).

## Discussion

ID and IDA remain serious global health concerns and current interventions, including iron fortification and supplementation, vary in their effectiveness as treatments for these conditions, in part due to unpleasant sensory changes or adverse side effects ^47^. Here we conducted a palatability and acceptability study to analyse sensory properties of fortified beverages containing different concentrations of either a mineral-enriched or a placebo powder. We found that the addition of either powder to tap water resulted in sensory changes, and that both the mineral-enriched powder and placebo were most palatable and acceptable to participants when reconstituted at a concentration of 1g/L (compared to 2g/L or 3g/L). Our study suggests that interventional products to combat ID and IDA can be formulated with improved consumer acceptability, which may help increase adherence when treating these conditions in women of reproductive age.

We demonstrate that fortification with a mineral-enriched powder containing electrolytic iron did not result in unpleasant sensory changes when reconstituted in tap water at the lowest concentration (1g/L). An inverse relationship was observed between the concentration of iron and hedonic scores for almost every sensory category, in both treatment and placebo groups. Specifically, the “odour”, “colour”, and “overall taste” (and thus, palatability) were the sensory properties most significantly affected by increased concentration. While to our knowledge there are no other studies that specifically investigate sensory changes to tap water fortified with electrolytic iron, our finding is consistent with results from similar studies involving other iron fortification systems ^48-52^. A series of studies that assessed the use of water fortified with ferrous sulphate for the prevention of IDA in children in Brazil found that 20 mg doses of iron were associated with more unfavourable sensory changes in the colour and taste of tap water; however, this was not observed when the water was fortified with lower amounts (10 mg) of iron ^49,53,54^. These findings suggest that fortification of tap water with lower iron dosages will be more favourable and palatable. Similarly, an investigation of the sensory properties (colour, odour, and taste) in cheese fortified with different amounts of ferrous sulphate (either 0.016 mg, 0.822 mg, or 0.932 mg of Fe/g of cheese) reported that the samples with the lowest amount of iron (0.016 mg) had the most favourable sensory properties across every category ^52^. Together, these findings demonstrate that fortification with lower doses of iron is consistently associated with improved palatability and favourability is important to consider in terms of consumer uptake and adherence, as the acceptability of commercial iron fortification programs can be negatively affected by unfavourable sensory changes to food vehicles, leading to poor adherence ^23,39,49,55^.

To investigate the acceptability of this mineral-enriched powder, we assessed the willingness of participants to drink the samples daily and found an inverse relationship between acceptability and sample concentration. In general, participants reported they would be less willing to drink the samples as the concentration of the active or placebo powder increased. This further suggests that our mineral-enriched powder will be more acceptable to the target population when reconstituted at lower concentrations, which may thus increase adherence to a treatment regimen using an iron fortified beverage ^56^. Although there are very few studies that investigate the acceptability of iron fortification systems in women of reproductive age, there have been several efficacy studies conducted in different populations that have reported participant adherence and acceptability as part of their secondary outcomes ^49,50,53,57^. Two research groups conducted randomised controlled trials in different paediatric populations to investigate the efficacy of food fortification using a micronutrient powder (“*Sprinkles*”) containing either 12.5, 20, or 30 mg of ferrous fumarate compared to supplementation with oral iron drops (“*DROPS*”) containing either 12.5 ^57^ or 20^53^ mg of ferrous sulphate ^53,57^. Both studies found that the serum ferritin levels in all participants increased after eight weeks of treatment, with no significant differences between groups; however, they did report that participant adherence to the consumption regimen varied significantly between treatment groups. Specifically, both studies found that participants randomised to low dose *Sprinkles* (12.5 mg of ferrous fumarate) had significantly better adherence rates than participants randomised to higher dose *Sprinkles* (20 or 30 mg of ferrous fumarate) or *DROPS* ^53,57^. These findings demonstrate that food fortification with low doses of iron can be both effective and acceptable to consumers, further supporting our results.

A strength of our study includes that the tested home-beverage fortification system used a mineral-enriched powder containing electrolytic iron, a relatively inert form of iron known to have sensory-related advantages over other common iron fortificants ^9,24,52,58,59^. While we recognise the importance of bioavailability when selecting an iron compound, the size of the iron particle is also an important consideration, as a smaller particle size help to increase bioabsorption^47,60^. The particle size of the electrolytic iron used in this study is very small (less than <20 μm) allowing us to focus on the sensory perceptions of this intervention in the current study in an effort to determine consumer acceptability. The World Health Organization also regards food fortification as one of the safest, most effective, and most affordable ways to administer dietary iron in deficient populations ^57,61^. However, iron is one of the most difficult micronutrients with which to fortify the diet, as it is known to interact strongly with many food constituents, resulting in unfavourable sensory changes to the colour, odour, and taste of fortified foods ^34,37,62^. Water soluble forms of iron (such as ferrous sulphate, ferrous gluconate, and ferrous fumarate) are most commonly used for oral iron replacement therapy given their high bioavailability; however, fortification with these iron compounds is frequently limited by their strong interactions with many foods ^38,58,62^. In contrast, fortification with electrolytic iron has been widely studied in many foods and has been consistently associated with significantly fewer organoleptic changes compared to water-soluble forms of iron fortified in the same foods ^48,51,63,64^. Researchers have even recommended that cereal staples be fortified with electrolytic iron when other water-soluble forms of iron result in unfavourable sensory changes ^47,58,65^. Given that the sensory perceptions of food can influence the likelihood of consumption ^41,52^, fortification with electrolytic iron can overcome the sensory barriers faced by treatment interventions involving water-soluble iron compounds, thus contributing to greater palatability and long-term consumer adherence ^34,47,59,61^. Further, electrolytic iron is more cost-effective than ferrous sulphate (the current leading compound in oral iron replacement therapy ^47^) which is advantageous, as economic feasibility is an important feature of sustainable treatments for micronutrient deficiencies ^47,66^. An additional strength of our study includes that we are one of the first to report on the palatability and acceptability of a fortification system involving electrolytic iron in women of reproductive age. Women are one of the most vulnerable populations at risk for ID and IDA ^11,14,17,67^, and thus there is a great need for palatable interventions that are accepted by this population for regular consumption of this product to effectively treat these conditions ^56^.

The generalizability of our results is limited by the small sample size and the subjective nature of the data collected. However, our approach used the 9-point hedonic scale, a validated methodological tool that has been used to assess the acceptability of foods and beverages for over 40 years ^68-71^. Further, the known organoleptic changes to food and beverages due to iron make the application of this scale an appropriate methodologic approach for our study ^34,61,66,72,73^.

We found that electrolytic iron fortification in tap water is palatable, acceptable, and favourable to women of reproductive age. These findings provide important preliminary data on the sensory perceptions of this preparation in one of the most vulnerable populations at risk for ID and IDA. Successful fortification systems consist of food or beverages that are already common in the diet and can thus be effectively incorporated into an individual’s daily routine ^61,74^. Evaluating consumer preferences for fortified foods and beverages, including palatability and acceptability, is also critical to inform the formulation or consumption regimen of these products ^46,75,76^, which will influence intervention success. Our findings suggest that electrolytic iron fortification of tap water could improve consumption of this intervention, and if efficacious in improving iron status, may ultimately help to strengthen global efforts to combat ID and IDA among women of reproductive age.

## Key Messages

- There is a need to develop effective and sustainable treatment interventions for ID and IDA in women of reproductive age, one of the populations most at risk for these conditions.
- Despite fortification being one of the most sustainable approaches to combat nutritional deficiencies, iron is one of the most difficult micronutrients to fortify as it can result in unacceptable sensory changes to fortified foods, ultimately leading to poor consumer adherence and decreased treatment efficacy.
- This study is one of the few to provide insight into the sensory perceptions of an iron fortification system in women of reproductive age, as previous studies on iron fortification systems have been largely focused on paediatric populations.
- The mineral enriched powder in this study, if also proven effective, supports global efforts towards developing sustainable treatments for ID and IDA in women of reproductive age.

## Data Availability

All data produced in the present work are contained in the manuscript.

## Acknowledgements

We would also like to acknowledge and thank the individuals who generously volunteered their time and data for the purposes of this study. Gum Products International Inc. (GPI) provided the proprietary food stabilizer in the mineral-enriched powder.

