## Supplemental Table 1 for "Sensory evaluations of a novel iron and zinc-enriched powder for the potential treatment and prevention of iron deficiency in women of reproductive age"

**Table 1.** Sensory properties and overall taste for active and placebo powders across concentrations

| Active Solutions | | | | |
|---|---|---|---|---|
| **Sensory Property of Samples** | **S1** (1 g/L) **(N=35)** | **S2** (2g/L) **(N=35)** | **S3** (3 g/L) **(N=35)** | **p-value** |
| Odour | 8 (6-9) [a] | 8 (5-9) [ab] | 6 (4-8) [c] | p<0.05 |
| Colour | 9 (8-9) [a] | 9 (8-9) [ab] | 8 (7-9) [c] | p<0.05 |
| Salty | 8 (6-9) [a] | 5 (3-8) [b] | 4 (3-6) [c] | p<0.05 |
| Sweet | 8 (7-9) [a] | 7 (6-9) [ab] | 6 (4-9) [c] | p=0.003 |
| Bitter | 8 (5-9) [a] | 5 (4-8) [b] | 4 (3-6) [c] | p<0.05 |
| Umami | 9 (6-9) | 8 (5-9) | 8 (4-9) | p=0.169 |
| Sour | 9 (8-9) [a] | 6 (4-9) [b] | 4 (3-5) [c] | p<0.05 |
| **Placebo Solutions** | | | | |
| **Sensory Property of Samples** | **S4 (**1 g/L) **(N=35)** | **S5** (2g/L) **(N=35)** | **S6** (3 g/L) **(N=35)** | **p-value** |
| Odour | 8 (5-9) [a] | 6 (4-8) [a] | 4 (3-6) [b] | p<0.05 |
| Colour | 8 (7-9) [a] | 6 (4-8) [b] | 4 (3-7) [c] | p<0.05 |
| Salty | 6 (4-7) [a] | 4 (2-5) [b] | 2 (2-4) [c] | p<0.05 |
| Sweet | 7 (6-9) [a] | 6 (4-7) [b] | 6 (3-8) [b] | p=0.003 |
| Bitter | 7 (4-9) [a] | 4 (2-5) [b] | 3 (2-5) [b] | p<0.05 |
| Umami | 7 (4-9) | 5 (4-7) | 4 (2-6) | p=0.07 |
| Sour | 7 (4-9) [a] | 4 (3-5) [b] | 3 (1-4) [c] | p<0.05 |

Hedonic scores of participants' (N=35) evaluation of each sensory property compared within groups for active samples (S1, S2, S3) and placebo samples (S4, S5, S6) at concentrations of 1g/L, 2g/L, and 3g/L. Hedonic scores range from 1-9, where a score of 1=Disliked extremely, 9=Liked extremely, and 5=Neither like nor dislike. Data are median (IQR). Groups with different letters are significantly different (Friedmans two-way analysis of variance by ranks test and Wilcoxon signed-rank test with Bonferroni correction for multiple comparisons, $p<0.05$).
