## Supplemental Table 2 for "Sensory evaluations of a novel iron and zinc-enriched powder for the potential treatment and prevention of iron deficiency in women of reproductive age"

**Table 2.** Proportion of participant scores for "overall taste" of active and placebo samples

| Hedonic Scores for Participant Liking | Proportion of Participants (n) | | |
|---|---|---|---|
| **Active Samples** | **S1** | **S2** | **S3** |
| Dislike (Scores 1-4) | 9% (3) | 34% (12) | 74% (26) |
| Neutral (Score of 5) | 9% (3) | 9% (3) | 11% (4) |
| Like (Scores 6-9) | 82% (29) | 57% (20) | 15% (5) |
| **Placebo Samples** | **S4** | **S5** | **S6** |
| Dislike (Scores 1-4) | 29% (10) | 74% (26) | 94% (33) |
| Neutral (Score of 5) | 11% (4) | 3% (1) | 0% (0) |
| Like (Scores 6-9) | 60% (21) | 23% (8) | 6% (2) |

Participants' (N=35) "overall liking" of each sample (S1-S6) using 9-point hedonic scale. Hedonic scores range from 1-9, where a score of 1=Disliked extremely, 9=Liked extremely, and 5=Neither like nor dislike. Active samples S1, S2, S3 contain the mineral-enriched powder at concentrations of 1g/L, 2g/L, and 3g/L, respectively. Placebo samples S4, S5, S6 contain the placebo powder at concentrations of 1g/L, 2g/L, and 3g/L, respectively. Proportion reporting overall disliking of each active and placebo sample include participants who gave hedonic scores between 1-4. Proportion reporting of overall liking of each active and placebo sample include participants who gave the samples hedonic scores between 6-9.
